# Personality profiles differ between patients with epileptic seizures and patients with psychogenic non-epileptic seizures

**DOI:** 10.1101/19002717

**Authors:** Michelle Leong, Albert D Wang, David Trainor, Ben Johnstone, Genevieve Rayner, Tomas Kalincik, Izanne Roos, Patrick Kwan, Terence J O’Brien, Dennis Velakoulis, Charles B Malpas

## Abstract

**Aim:** The primary aim of the study was to determine whether patients with psychogenic non-epileptic seizures (PNES) have different personality profiles compared to patients with epileptic seizures (ES). The secondary aim was to determine whether any such personality differences could be used to efficiently screen for PNES in clinical settings.

**Background:** PNES and ES are often difficult to differentiate, leading to incorrect or delayed diagnosis. While the current gold-standard investigation is video-EEG monitoring (VEM), it is resource intensive and not universally available. Although some research has investigated the differential psychological profiles of PNES and ES patients, most studies have focused on symptoms of psychopathology. The lack of research using modern personality models in PNES and ES presents a gap in knowledge that this study aimed to address.

**Methods:** A retrospective collection of data was conducted on patients who completed the NEO-Five Factor Inventory questionnaire during a VEM admission to the Royal Melbourne Hospital between 2002-2017. Patients were classified as either ES or PNES based on clinical consensus diagnosis. For patients with ES, type of epilepsy and laterality of seizure focus were also recorded. Personality differences were investigated using Bayesian linear mixed effects models. Receiver operating characteristic curve analysis was also performed to generate sensitivities and specificities of individual personality scores.

**Results:** 305 patients were included in the study. The ‘openness to experience’ domain was the only personality factor demonstrating strong evidence for a group difference (BF_10_ = 21.55, *d* = −0.43 [95% CI −0.71, −0.17]), with patients in the PNES group having higher scores compared to the ES group. Within the openness to experience domain, only the ‘aesthetic interest’ facet showed evidence for a group difference (BF_10_ = 7.98, *d* = −0.39 [95% CI −0.66, −0.12]). ES patients had lower scores on these measures compared to the normal population, while PNES patients did not. Both openness to experience and aesthetic interest, however, showed poor sensitivities (53%, 46% respectively) and specificities (69%, 46% respectively) for classifying PNES and ES patients. There were no differences between personality profiles in Temporal Lobe Epilepsy (TLE) and non-TLE patients, or in laterality in TLE.

**Conclusion:** Patients with ES exhibit lower openness to experience and aesthetic interest compared to patients with PNES and compared to the general population. Despite these differences, the relatively low sensitivity and specificity of these instruments suggests their use is limited in a clinical setting. Nevertheless, these findings open up new avenues of research using modern personality models to further understand patients with epilepsy and related presentations.

## 1. Introduction

Psychogenic non-epileptic seizures (PNES) are episodes of abnormal limb movements that resemble epileptic seizures (ES), but are not associated with organic aetiologies, and presumably manifest due to psychological distress [1]. Similarities in presentation to ES mean that patients with PNES are often misdiagnosed with epilepsy, resulting in multiple financial, social, emotional and health implications [2–6]. The current gold standard investigation in the differentiation between ES and PNES is video-EEG monitoring (VEM) [7–10]. Despite VEM’s advantages, it is expensive, of limited availability and resource intensive [11]. The cost of VEM approximates to USD $1,100-$1,700 per day due to the need for a highly trained multidisciplinary team and specialised equipment [11,12]. This presents a challenge for socioeconomically disadvantaged populations. Furthermore, the occurrence of a PNES event is not guaranteed, with PNES being more likely to occur off-camera than ES [13]. Therefore, a pressing issue for PNES is to find a simpler, cheaper and more readily available methods to screen for PNES.

Due to this recognizable problem, there is a need to better identify patients at risk of PNES prior to being admitted for VEM [14]. A notable area of research has been the differential psychological profiles of PNES and ES patients. Most research in this area has focused on measures of psychopathology, such as the Minnesota Multiphasic Personality Inventory (MMPI) and the Personality Assessment Inventory (PAI). While these measures assess some aspects of personality, they focus on common symptoms of psychiatric disorders (such as depression, or anxiety). For example, while the MMPI does include the assessment of some personality characteristics, the psychopathological variables make it hard to determine personality correlations between patients with ES and PNES. Furthermore, the PAI’s clinical scales have been derived based on DSM criteria [15]. Relatively little research has used modern models of non-pathological personality traits such as the five-factor model (FFM). The FFM is a statistically-derived model that describes personality along five domains, including neuroticism, extraversion, openness to experience, agreeableness, and conscientiousness [16].

Few notable studies using the five-factor model in the differentiation of PNES and ES has been performed thus far. In the process of cluster analysing PNES using the Revised NEO Personality Inventory (NEO PI-R), Cragar et al. [17] compared the PNES patients to an epilepsy control group and found that PNES patients scored higher on neuroticism and on the agreeableness facet of modesty and lower on the extraversion facet of gregariousness and on the agreeableness facet of trust compared to epilepsy controls. Although these findings are relevant, the study sizes were small and personality differences between PNES and epilepsy were not the focus of the study, but rather a secondary finding. More recently, Vilyte et al. reported no differences in FFM traits between patients with PNES and ES in a small sample of South African patients [18]. Lack of clinically focused, high-quality research in this area shows the need for a more in-depth study using the modern personality models.

This large retrospective study aimed to utilise the FFM model of personality to determine profile differences between patients with PNES and ES admitted to a specialist VEM unit in a large tertiary centre. It also aimed to directly investigate the classification accuracy of these differences in a clinical setting, which has often been overlooked in previous research. Following Cragar et al., we hypothesised that PNES patients would score higher on neuroticism, and lower on extraversion compared to patients with ES. We expected that such differences would translate into efficient screening for PNES in this population.

## 2. Methods

### 2.1 Participants

This was a retrospective case control study. Data were collected from patients as part of routine psychological care who were admitted to the Royal Melbourne Hospital (RMH), Australia, VEM unit between the years 2002-2017. The RMH is a major epilepsy centre in Australia and utilises the use of VEM and a multidisciplinary team involving epileptologists, psychiatrists, EEG scientists, and neuropsychologists in diagnosing epilepsy. Inclusion criteria for the current study included patients who had completed the NEO Five-Factor Inventory (NEO-FFI) questionnaire during their VEM admission and those who had a 6^th^ grade reading level of English and above. Exclusion criteria included patients who were under the age of 17 and patients with 10 or more responses missing. Of the 344 patients who were initially recruited, 8 patients were underage and 31 had missing responses, leading to 305 patients being finally included in the study. This study received approval from the Melbourne Health Ethics Committee (MH HREC# QA2012044).

### 2.2 Clinical diagnosis

Diagnosis of the patients admitted for VEM was determined at a large multidisciplinary meeting consisting of EEG scientists, epileptologists, neuropsychologists, and psychiatrists. Neuropsychiatric assessments, neurological examinations, and imaging was also heavily considered in making the final diagnosis. A diagnosis of Epilepsy was made when a patient met the International League Against Epilepsy (ILAE) criteria of epilepsy [19,20], while a diagnosis of PNES was made based on abnormal movements in the absence of EEG changes [21]. Patients were classified into 4 main categories: ES, PNES, ES+PNES and non-diagnostic. Patients were categorised as non-diagnostic when a definite diagnosis could not be made or if they had a condition which did not meet either ES or PNES criteria. Conditions reported were migraines, postural hypotension, anxiety, cardiac syncope, arrhythmia, alcohol excess, post-traumatic stress disorder (PTSD), benign paroxysmal positional vertigo (BPPV) and sick sinus syndrome. For patients with ES, the type of epilepsy was also classified according to the ILAE classification of epilepsies [22]. Specifically, the focal epilepsies, temporal lobe epilepsy (TLE) and extra-temporal lobe epilepsy (extra-TLE), and generalised epilepsy. Laterality of seizure focus was also recorded for TLE patients. The NEO-FFI results were not known to the VEM team and did not inform the diagnosis of PNES.

### 2.3 Personality assessment

The NEO-FFI is a 60-item self-administered measure of personality that describe an individual’s degree of standing across five higher order factors: neuroticism, extraversion, openness to experience, agreeableness, and conscientiousness [16]. These domains and their definitions are included in *Table A.1* in the supplementary material. The five factors were identified via the factor analysis of all the trait adjectives found in English and other natural languages [23] and has received widespread empirical support [16,24,25]. Construct validity and internal consistency studies have shown the NEO-FFI to be a valid and reliable psychometric tool for the assessment of personality [16]. Furthermore, the five-factor model has shown to have good cross-cultural replicability [26] and real-life applicability by demonstrating its ability to predict numerous important life outcomes [16]. The NEO-FFI scores are calculated in the form of a normalised score, providing an indication as to where an individual lies on a spectrum compared to the general population [16].

Facet scores were also computed for each of the 5 domains. While domain scores allow for a global picture of an individual’s personality, facets allow for a deeper understanding of the individual [16]. We computed scores for the 13 facets derived via an analysis performed by Saucier [27]. *Table A.2* illustrates the domains and its facets.

### 2.4 Statistical analysis

All analyses were performed JASP [28] and R [29]. Bayesian linear mixed effects models were used to determine differences in personality profiles between the PNES and ES groups. Separate analyses were conducted for TLE and non-TLE groups, as well as laterality in TLE patients. For all analyses, the null model included gender and age as predictors of personality and the alternative model contained these predictors plus the relevant personality domains and diagnostic group (e.g. PNES vs ES). The statistical importance of each term was determined using a model averaging approach. Specifically, the Bayes factor of inclusion (BF_inc_) was computed for each term which represents the sum of the posterior model probabilities for all models containing the term divided by the sum of posterior probabilities for all models not containing the term. Higher interactions are excluded. A BF_inc_ > 3 was taken to indicate evidence for an effect, while BF_inc_ < 1/3 was taken as evidence for the null hypothesis. BFs between 1/3 and 3 were taken to indicate insensitivity to either hypothesis. Follow-up Bayesian independent sample T-tests were computed to identify specific domain differences between the groups, with BFs interpreted as described above. Effect sizes were reported for all group comparisons in the form of Cohen’s [30] *d*, with 95% credible intervals. Receiver operating characteristic (ROC) curve were computed to generate sensitivities and specificities of individual domains and facets in order to determine usefulness in a clinical setting.

## 3. Results

### 3.1 Sample characteristics

Full diagnosis and demographic variables are described in *Table 1*. Of the 305 patients who met eligibility criteria, 122 (40%) were diagnosed with ES, 14 (5%) with both ES and PNES, and 90 (30%) with PNES. The remaining 79 (25%) patients were classified as either ‘non-diagnostic’, or were diagnosed with an alternative, non-epileptic or non-psychogenic cause of their presentation. Only the ES and PNES patients were included in the main statistical analyses. Of the ES patients, 23 (17%) had extra-temporal focal epilepsy, 74 (54%) had TLE, 31 (23%) had generalised epilepsy, and 8 (6%) had probable focal epilepsy of unspecified or unclear focus. Mean age was 38.79 (SD 15.33, range 17-92) and the majority of participants were female (*n*= 202, 66%). There was no relationship between gender and VEM diagnosis (BF_10_ = 0.02).

**Table 1.**
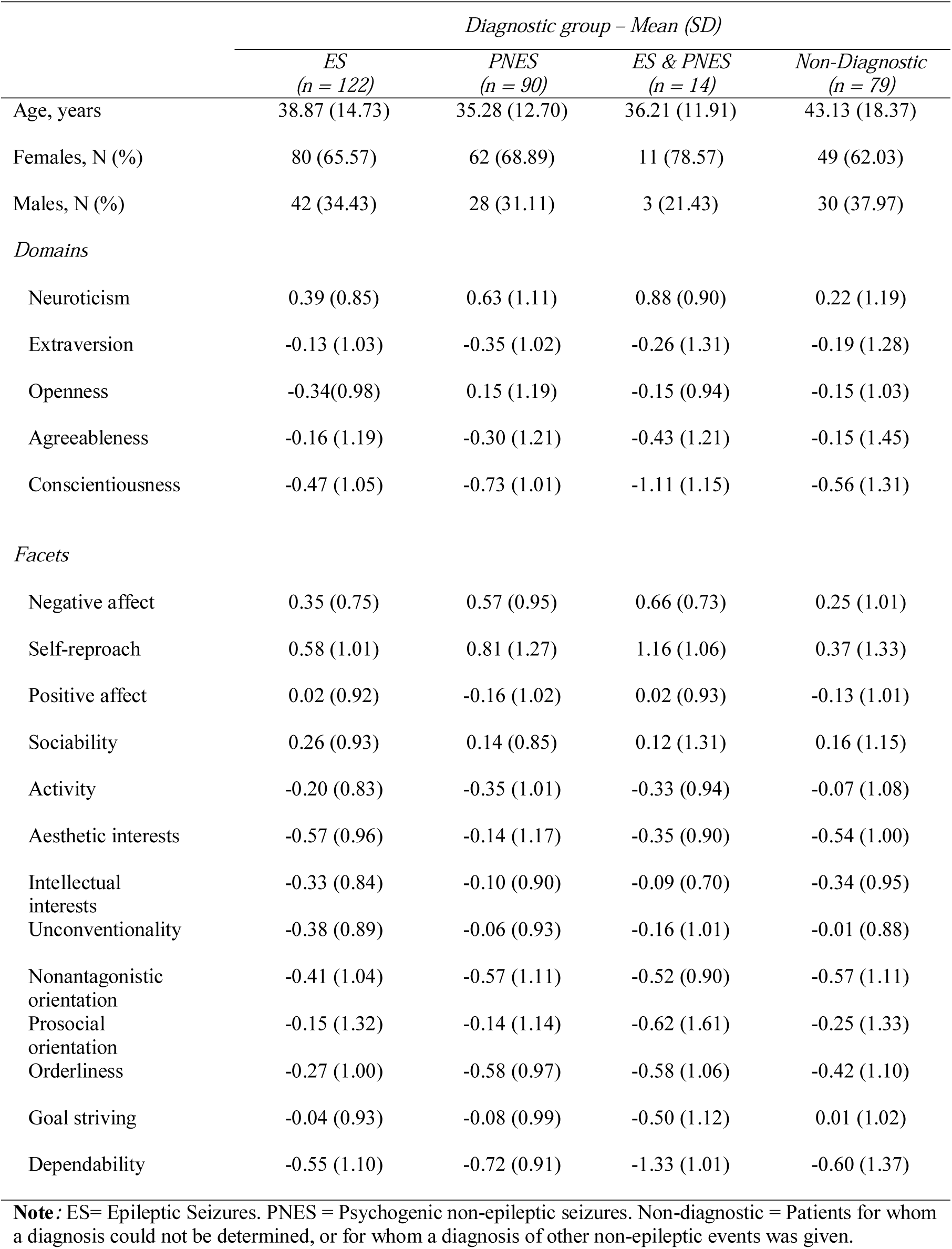
Sample characteristics

|  | *Diagnostic group – Mean (SD)* |  |  |  |
| --- | --- | --- | --- | --- |
|  | *ES*<br>(*n* = 122) | *PNES*<br>(*n* = 90) | *ES & PNES*<br>(*n* = 14) | *Non-Diagnostic*<br>(*n* = 79) |
| Age, years | 38.87 (14.73) | 35.28 (12.70) | 36.21 (11.91) | 43.13 (18.37) |
| Females, N (%) | 80 (65.57) | 62 (68.89) | 11 (78.57) | 49 (62.03) |
| Males, N (%) | 42 (34.43) | 28 (31.11) | 3 (21.43) | 30 (37.97) |
| *Domains* |  |  |  |  |
| Neuroticism | 0.39 (0.85) | 0.63 (1.11) | 0.88 (0.90) | 0.22 (1.19) |
| Extraversion | -0.13 (1.03) | -0.35 (1.02) | -0.26 (1.31) | -0.19 (1.28) |
| Openness | -0.34(0.98) | 0.15 (1.19) | -0.15 (0.94) | -0.15 (1.03) |
| Agreeableness | -0.16 (1.19) | -0.30 (1.21) | -0.43 (1.21) | -0.15 (1.45) |
| Conscientiousness | -0.47 (1.05) | -0.73 (1.01) | -1.11 (1.15) | -0.56 (1.31) |
| *Facets* |  |  |  |  |
| Negative affect | 0.35 (0.75) | 0.57 (0.95) | 0.66 (0.73) | 0.25 (1.01) |
| Self-reproach | 0.58 (1.01) | 0.81 (1.27) | 1.16 (1.06) | 0.37 (1.33) |
| Positive affect | 0.02 (0.92) | -0.16 (1.02) | 0.02 (0.93) | -0.13 (1.01) |
| Sociability | 0.26 (0.93) | 0.14 (0.85) | 0.12 (1.31) | 0.16 (1.15) |
| Activity | -0.20 (0.83) | -0.35 (1.01) | -0.33 (0.94) | -0.07 (1.08) |
| Aesthetic interests | -0.57 (0.96) | -0.14 (1.17) | -0.35 (0.90) | -0.54 (1.00) |
| Intellectual interests | -0.33 (0.84) | -0.10 (0.90) | -0.09 (0.70) | -0.34 (0.95) |
| Unconventionality | -0.38 (0.89) | -0.06 (0.93) | -0.16 (1.01) | -0.01 (0.88) |
| Nonantagonistic orientation | -0.41 (1.04) | -0.57 (1.11) | -0.52 (0.90) | -0.57 (1.11) |
| Prosocial orientation | -0.15 (1.32) | -0.14 (1.14) | -0.62 (1.61) | -0.25 (1.33) |
| Orderliness | -0.27 (1.00) | -0.58 (0.97) | -0.58 (1.06) | -0.42 (1.10) |
| Goal striving | -0.04 (0.93) | -0.08 (0.99) | -0.50 (1.12) | 0.01 (1.02) |
| Dependability | -0.55 (1.10) | -0.72 (0.91) | -1.33 (1.01) | -0.60 (1.37) |

**Note:** ES= Epileptic Seizures. PNES = Psychogenic non-epileptic seizures. Non-diagnostic = Patients for whom a diagnosis could not be determined, or for whom a diagnosis of other non-epileptic events was given.

Psychiatric comorbidity was common with 113 (37%) patients having a history of any major psychiatric disorder, and 12 (4%) patients having a history of borderline personality disorder. PNES participants were more likely to have a history of depression (BF_10_ = 4.66), current depression (BF_10_=5.91), or any current major psychiatric disorder (BF_10_ = 16.95). There was no evidence for a relationship between diagnostic category and history of any major psychiatric disorder (BF_10_ = 1.52), history of anxiety disorder (BF_10_ = 2.23), current anxiety disorder (BF_10_ = 1.36), or history of borderline personality disorder (BF_10_ = 2.87).

### 3.2 Comparison of PNES and ES patients to population normative data

A Bayesian liner mixed effects model was computed to determine whether there were differences between personality domains and facets in the combined PNES and ES cohort. There was strong evidence for an effect of domain (BF_inc_ > 100). As shown in *Figure 1*, the highest scores were observed for neuroticism (*M* = 0.49, *SD* = 0.97), followed by openness (*M* = 10.13, SD = 1.10), agreeableness (*M* = −0.22, *SD* = 1.20), extraversion (*M* = −0.23, *SD* = 1.03), and conscientiousness (*M* = −0.58, *SD* = 1.04). There was strong evidence that the levels of neuroticism were higher when compared to normal population (BF_10_ ≥ 100). Levels of extraversion (BF_10_ = 10.84) and consciousness (BF_10_ ≥100) were significantly lower than the normal population.

**Figure 1.**
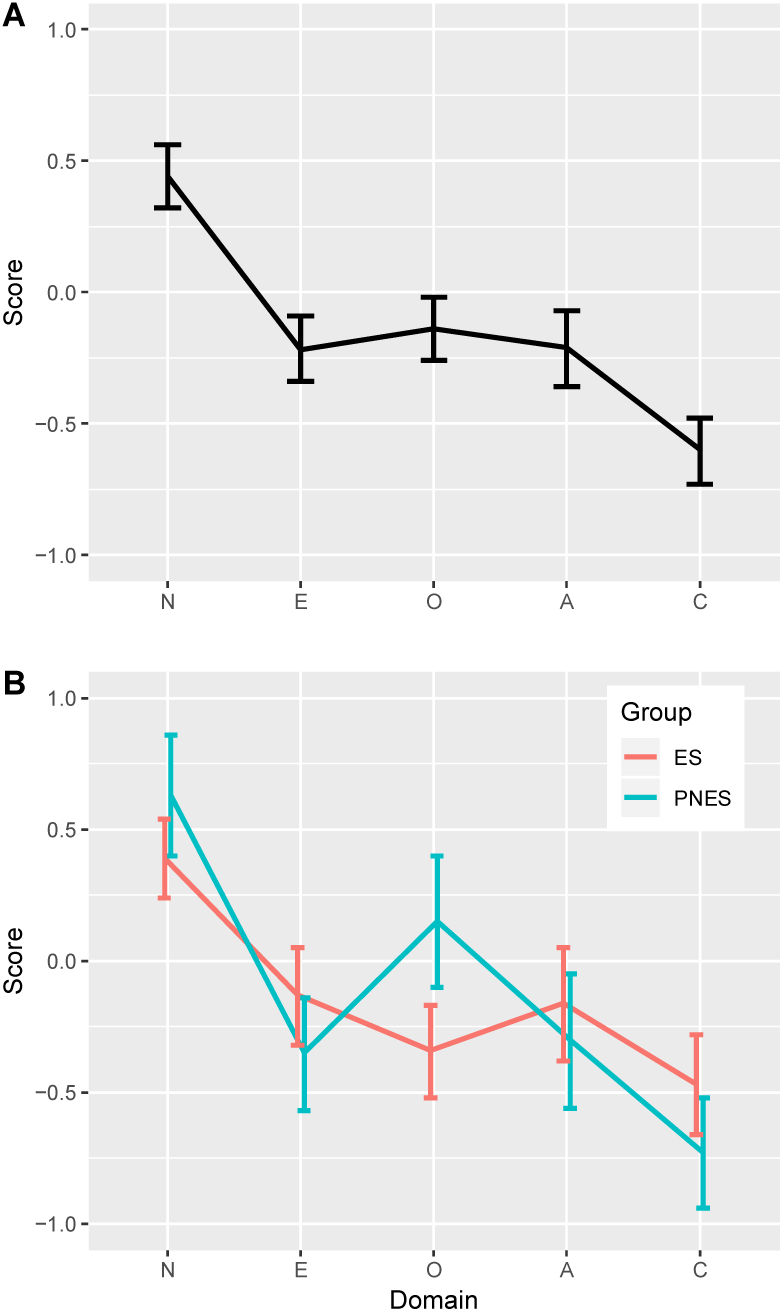
Distribution of personality scores in the combined PNES and ES cohort. The dotted line shows the mean for the general population. As shown in **A**, participants had higher scores on neuroticism, and lower scores in extraversion and conscientiousness compared to the general population. As shown in **B**, the two groups had comparable scores for all domains except openness to experience, on which the PNES group had high scores compared to the ES group. Scores for the ES group were below the mean for the general population.

**Figure 2.**
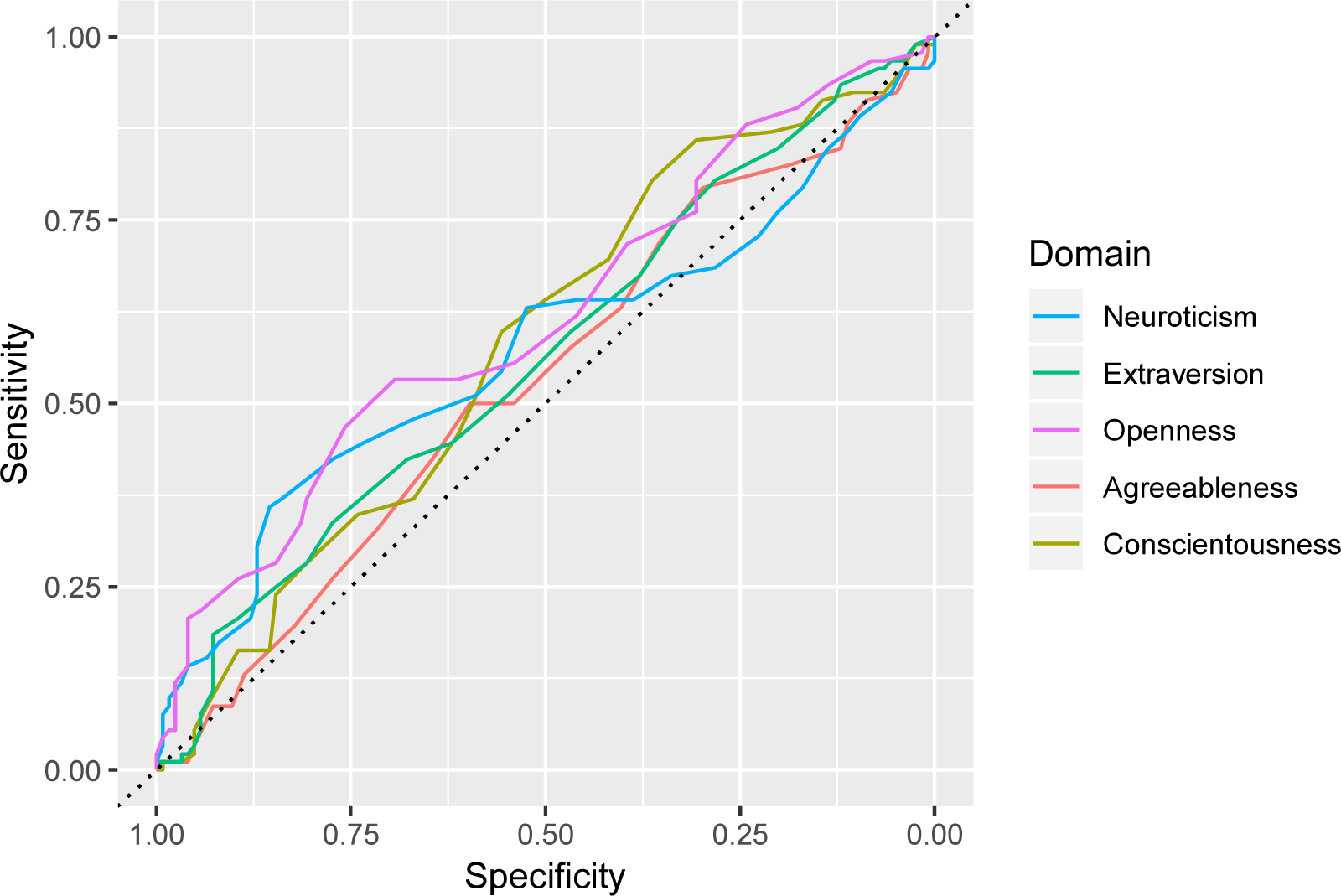
Receiver operating characteristic (ROC) curves for personality score domains in terms of classification of PNES and ES.

**Figure 3.**
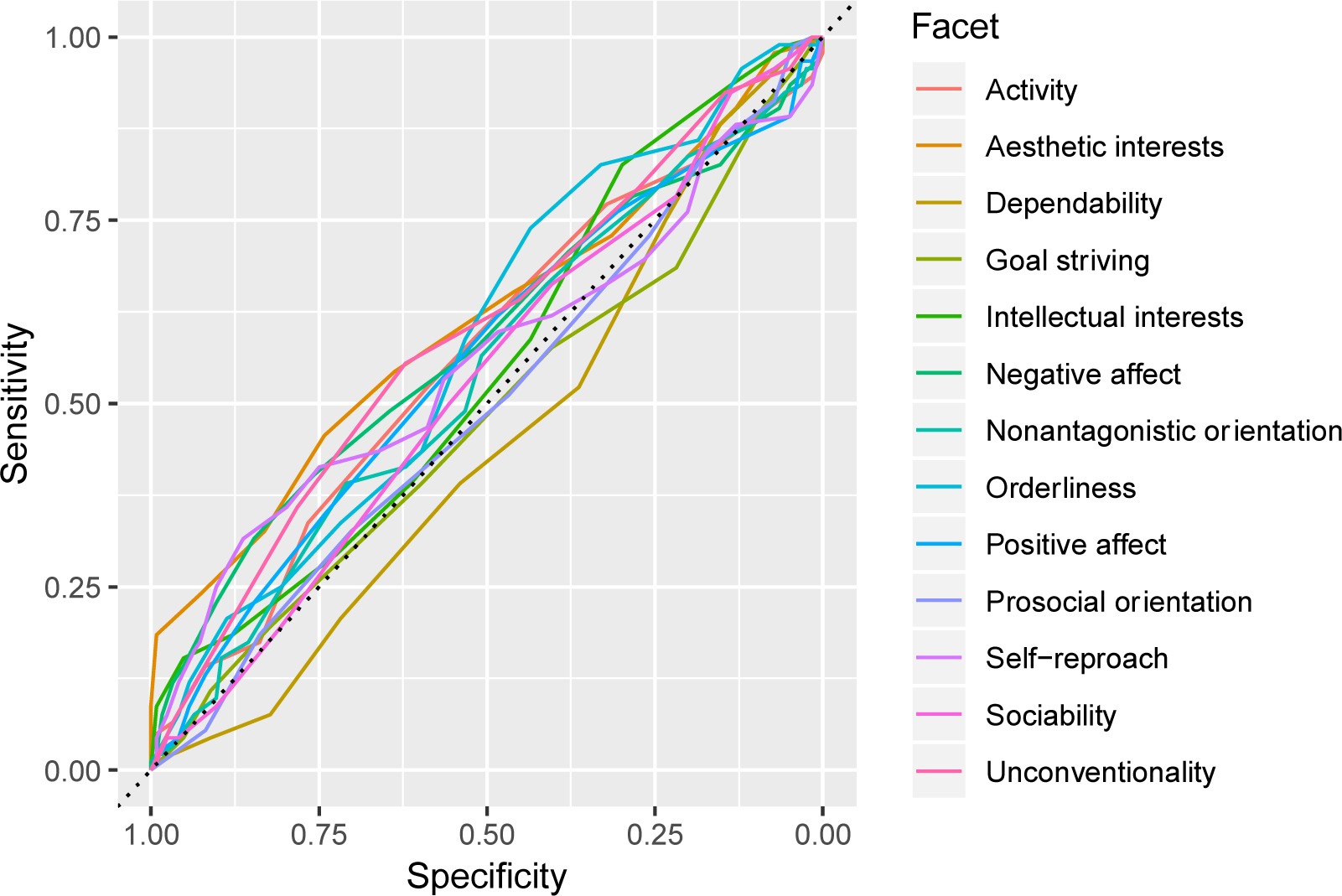
Receiver operating characteristic (ROC) curves for personality score facets in terms of classification of PNES and ES.

### 3.3 Comparison of Epilepsy and PNES groups

A Bayesian linear mixed effects model was computed to determine differences in personality domains between diagnostic groups (PNES and ES). While there was strong evidence for a main effect of domain (BF_inc_ >100), there was no evidence for a main effect of diagnostic group (BF_inc_ = 1.86). There was, however, evidence for diagnostic group by domain interaction (BF_inc_ = 9.92). This indicates that the two diagnostic groups had different personality profiles. Follow-up Bayesian independent samples T-tests were performed to investigate which specific domains differed between groups (see *Table 2* and *Figure 1*). Openness to experience was the only domain found with a strong evidence for a group difference (BF_10_ = 21.55, *d* = −0.43 [95% CI −0.71, −0.17]). Patients in the PNES group reported greater openness to experience compared to patients in the ES group. The null hypothesis was supported for the agreeableness domain (BF_10_ = 0.21, *d* = 0.11 [95% CI −0.16, 0.38]), while the evidence was insensitive for the domains of neuroticism, extraversion and conscientiousness.

**Table 2.**
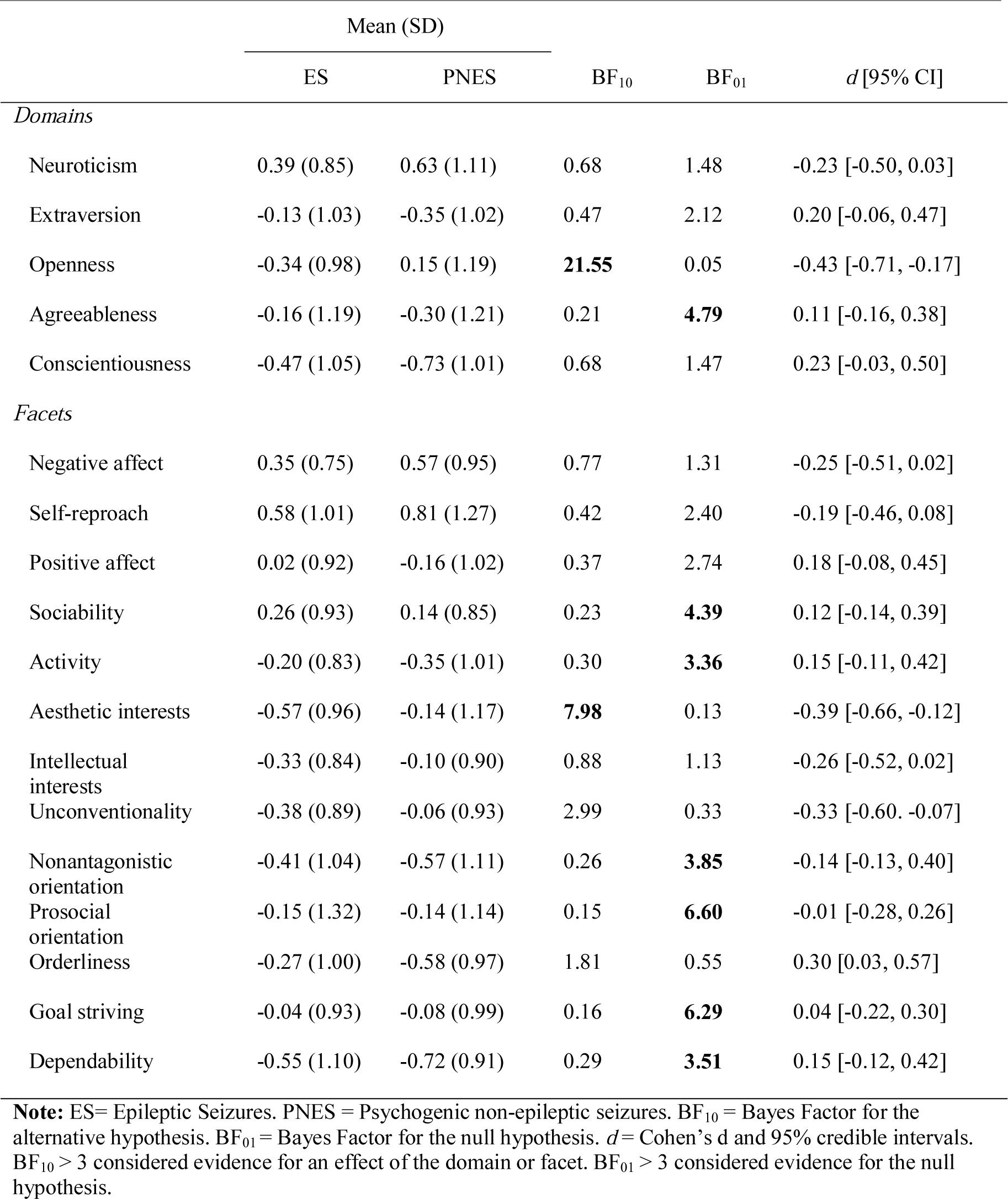
Comparison between diagnostic groups (PNES and ES)

|  | ES | PNES | BF<sub>10</sub> | BF<sub>01</sub> | *d* [95% CI] |
| --- | --- | --- | --- | --- | --- |
| *Domains* |  |  |  |  |  |
| Neuroticism | 0.39 (0.85) | 0.63 (1.11) | 0.68 | 1.48 | -0.23 [-0.50, 0.03] |
| Extraversion | -0.13 (1.03) | -0.35 (1.02) | 0.47 | 2.12 | 0.20 [-0.06, 0.47] |
| Openness | -0.34 (0.98) | 0.15 (1.19) | **21.55** | 0.05 | -0.43 [-0.71, -0.17] |
| Agreeableness | -0.16 (1.19) | -0.30 (1.21) | 0.21 | **4.79** | 0.11 [-0.16, 0.38] |
| Conscientiousness | -0.47 (1.05) | -0.73 (1.01) | 0.68 | 1.47 | 0.23 [-0.03, 0.50] |
| *Facets* |  |  |  |  |  |
| Negative affect | 0.35 (0.75) | 0.57 (0.95) | 0.77 | 1.31 | -0.25 [-0.51, 0.02] |
| Self-reproach | 0.58 (1.01) | 0.81 (1.27) | 0.42 | 2.40 | -0.19 [-0.46, 0.08] |
| Positive affect | 0.02 (0.92) | -0.16 (1.02) | 0.37 | 2.74 | 0.18 [-0.08, 0.45] |
| Sociability | 0.26 (0.93) | 0.14 (0.85) | 0.23 | **4.39** | 0.12 [-0.14, 0.39] |
| Activity | -0.20 (0.83) | -0.35 (1.01) | 0.30 | **3.36** | 0.15 [-0.11, 0.42] |
| Aesthetic interests | -0.57 (0.96) | -0.14 (1.17) | **7.98** | 0.13 | -0.39 [-0.66, -0.12] |
| Intellectual interests | -0.33 (0.84) | -0.10 (0.90) | 0.88 | 1.13 | -0.26 [-0.52, 0.02] |
| Unconventionality | -0.38 (0.89) | -0.06 (0.93) | 2.99 | 0.33 | -0.33 [-0.60, -0.07] |
| Nonantagonistic orientation | -0.41 (1.04) | -0.57 (1.11) | 0.26 | **3.85** | -0.14 [-0.13, 0.40] |
| Prosocial orientation | -0.15 (1.32) | -0.14 (1.14) | 0.15 | **6.60** | -0.01 [-0.28, 0.26] |
| Orderliness | -0.27 (1.00) | -0.58 (0.97) | 1.81 | 0.55 | 0.30 [0.03, 0.57] |
| Goal striving | -0.04 (0.93) | -0.08 (0.99) | 0.16 | **6.29** | 0.04 [-0.22, 0.30] |
| Dependability | -0.55 (1.10) | -0.72 (0.91) | 0.29 | **3.51** | 0.15 [-0.12, 0.42] |

**Note:** ES= Epileptic Seizures. PNES = Psychogenic non-epileptic seizures. BF<sub>10</sub> = Bayes Factor for the alternative hypothesis. BF<sub>01</sub> = Bayes Factor for the null hypothesis. *d* = Cohen's *d* and 95% credible intervals. BF<sub>10</sub> > 3 considered evidence for an effect of the domain or facet. BF<sub>01</sub> > 3 considered evidence for the null hypothesis.

A focused analysis was performed on the three facets that comprise the openness domain (see *Table 2*). There was evidence for group differences on the aesthetic interest facet (BF_10_ = 7.98, *d* = −0.39 [95% CI −0.66, −0.12]), with PNES patients reporting higher scores compared to ES patients. The evidence for the facets of intellectual interests and unconventionality was insensitive. No evidence for a group difference was observed for the remaining facets, and the null hypothesis was supported for the facets of sociability, activity, nonantagonistic orientation, prosocial orientation, goal striving, and dependability.

To investigate these findings further, Bayesian single sample T test were performed to compare each group to the general population in terms of levels of openness to experience. Patients in the ES group reported lower openness to experience (BF_10_ = 94.07, *d* = −0.34 [95% CI −0.52, −0.160]) and aesthetic interests (BF_10_ > 100, *d* = −0.58 [95% CI −0.77, −0.39]). In contrast, patients in the PNES group had scores on openness to experience (BF_10_ = 0.23, *d* = 0.12 [95% CI −0.09, 0.32]) and aesthetic interests (BF_10_= 0.21, *d* = −0.11, [95% CI −0.32,0.09]) that were not different to the general population.

### 3.4 Comparison of all diagnostic groups

A Bayesian linear mixed effects model was also computed to determine differences in personality domains between all four diagnostic groups (PNES, ES, ES+ PNES and non-diagnostic). While there was strong evidence for a main effect of domain (BF_inc_>100), the null hypothesis was supported for the main effect of diagnostic group (BF_inc_ < .01). The null hypothesis was also supported for the domain by diagnostic group interaction (BF_inc_ = 0.02). Examination of the means for these groups confirmed that the addition of the ES+PNES and non-diagnostic groups diluted the statistical effects.

### 3.5 Comparison of TLE and non-TLE

A Bayesian linear mixed effects model was computed to determine differences in personality domains between the ES patients with TLE and the non-TLE ES patients. There was strong evidence for a main effect of domain (BF_inc_ > 100). The null hypothesis was supported for the main effect of group (TLE vs non-TLE; BF_inc_ = 0.11) and the group by domain interaction (BF_inc_ = 0.10).

### 3.6 Comparison of Left and Right TLE patients

Of the 70 TLE patients, 3 patients with bilateral TLE were excluded from the analysis, leaving 37 Left TLE and 26 Right TLE patients. A Bayesian linear mixed effects model was computed to determine differences in personality domains between left and right TLE patients. The evidence was insensitive to the main effect of domain in this smaller sub-grouping (BF_inc_ = 2.36). The null hypothesis was support for the main effect of laterality (BF_inc_ = 0.17) and the laterality by domain interaction (BF_inc_ = 0.05).

### 3.7 Diagnostic accuracy

Receiver operating characteristic (ROC) curves were computed to determine whether personality scores accurately classified PNES and ES patients. As shown in *Table 3*, the openness to experience domain had an area under the curve (AUC) greater than .50, which suggests that it performs better than chance at classifying diagnostic group. The sensitivity (53%) and specificity (69%), however, were low. Similar results were seen for the aesthetic interests facet in terms of sensitivity (46%) and specificity (74%). None of the other personality domains produced high sensitivity or specificity metrics.

**Table 3.** Classification performance for personality domain and facet scores

| *Psychometric marker* | *AUC [95%CIs]* | *Threshold* | *Sensitivity* | *Specificity* | *PPV* | *NPV* |
| --- | --- | --- | --- | --- | --- | --- |
| *Domains* |  |  |  |  |  |  |
| Neuroticism | .57 [.49, .65] | > 1.20 | .36 | .85 | .65 | .64 |
| Extaversion | .56 [.49, .64] | < -1.40 | .18 | .93 | .65 | .61 |
| Openness | **.62 [.54, .69]** | > 0.08 | .53 | .69 | .56 | .67 |
| Agreeableness | .53 [.45, .61] | < -0.40 | .50 | .60 | .48 | .62 |
| Conscientiousness | .59 [.50, .66] | < -0.01 | .80 | .36 | .48 | .71 |
| *Facets* |  |  |  |  |  |  |
| Negative affect | .58 [.50, .66] | > 1.08 | .32 | .85 | .60 | .62 |
| Self-reproach | .55 [.48, .64] | > 1.66 | .32 | .86 | .63 | .63 |
| Positive affect | .56 [.48, .63] | < 0.19 | .62 | .48 | .47 | .63 |
| Sociability | .53 [.45, .61] | < 0.51 | .66 | .40 | .45 | .62 |
| Activity | .56 [.48, .64] | < -0.41 | .52 | .59 | .48 | .62 |
| Aesthetic interests | **.61 [.53, .69]** | > 0.10 | .46 | .74 | .57 | .65 |
| Intellectual interests | .55 [.48, .63] | > -0.87 | .83 | .30 | .47 | .70 |
| Unconventionality | **.59 [.52, .69]** | > -0.13 | .55 | .62 | .52 | .65 |
| Nonantagonistic orientation | .54 [.46, .62] | < -1.02 | .39 | .71 | .50 | .61 |
| Prosocial orientation | .50 [.42, .58] | > -3.02 | .99 | .04 | .43 | .83 |
| Orderliness | **.58 [.51, .66]** | < -0.01 | .74 | .44 | .49 | .69 |
| Goal striving | .48 [.40, .56] | > 1.19 | .11 | .91 | .48 | .58 |
| Dependability | .44 [.37, .52] | > -1.79 | .88 | .15 | .44 | .63 |

**Note:** AUC = area under the curve. CIs = 95% credible intervals. Threshold is the optimal cut-off for classifying PNES patients. PPV = positive predictive value. NPV = negative predictive value.

## 4. Discussion

In this study, we investigated differences in personality profiles between patients with PNES and ES using the five-factor model of personality. Our primary finding was that patients with PNES and ES differed in their personality profiles. Specifically, patients in the PNES group exhibited higher scores on the openness to experience domain, which measures such personality facets as propensity for fantasy, aesthetics interests, openness to feelings, and acceptance of new ideas. Follow-up analyses showed that the strongest evidence was for the aesthetic interests facet. Patients with ES had lower scores on these variables compared to the general population, while PNES patients did not differ from normative data. While these differences were large and supported by strong statistical evidence, the sensitivity and specificity for the openness and aesthetic interests scores were poor, which suggests that their use as psychometric screening instruments for PNES might be limited in a clinical setting. We also found that, when combined, the PNES and ES groups had higher levels of neuroticism, and lower levels of both extraversion and conscientiousness compared to the general population. Further analysis of the data revealed no differences between personality profiles in TLE and non-TLE patients, and no differences in personality profiles between patients with left or right TLE.

Our finding of elevation in openness and aesthetic interests in PNES patients is not entirely consistent with the previous literature. In a secondary analysis performed by Cragar et al., PNES patients scored higher in the domain of neuroticism compared to ES patients, while differences in the domain of openness were also found to be insignificant [17]. This discrepancy could be accounted for by several factors. One potential factor is the use of the NEO PI-R that was used in Cragar’s study as opposed to the NEO-FFI. The NEO PI-R assessment allows for personality to be described across 30 facet scales in addition to the 5 domains [16]. This may allow for more comprehensive coverage of personality, accounting for differences in results. However, the NEO-FFI was used in our study due to time constraints and has been proven to be both a reliable and adequate measurement of a global assessment of personality - the main focus of our study [16]. Results for neuroticism, conscientiousness and extraversion were also insensitive in our study. It is possible that our statistical power may have been insufficient to determine the results for these specific domains. However, if that it is true, the effect sizes are likely to be small and possibly of little clinical significance. Nevertheless, the finding of an elevation of openness is novel and adds to the body of research reinforcing the association between personality profiles and diagnostic groups.

One possible explanation for the elevation of openness and aesthetic interests may be related to the association of both openness and ES with cognition. Openness is one of the more challenging of the five personality factors to describe, due to the wide variety of traits that it seems to encompass [31]. There has been a great deal of contention in the literature regarding naming this specific factor, with other contenders being intellect and culture [31]. McCrae and Costa who coined the term openness described it as essentiality traits relating to the exploration of the world from a cognitive point of view [31]. It is, therefore, the factor that has the greatest correlation with cognitive ability amongst the five factors of personality[31,32]. Particularly in cognitive domains such as working memory, latent inhibition and implicit learning [31]. Epilepsy also has been known to have a strong association with cognition, with chronic epileptic patients having higher degrees of cognitive impairment [33]. There is evidence that patients who have poor seizure control, longer seizure duration, and earlier onset of disease are more adversely affected cognitively [33]. Given that this is most representative of our VEM cohort, it is possible that observed lower openness scores might be partly reflective of prevalent cognitive impairment in this group. However, it is important to note that this finding is speculative and provides a clear direction for future research.

The idea of a personality profile specific to TLE, the Geschwind syndrome, has had a long history. According to Waxman and Geschwind these patients demonstrate placidity, flighty attention span, hypersexuality, hypergraphia and hyperreligiousity [34–37]. This discovery led to heightened interests surrounding TLE personality and the development of the Bear-Fedio Inventory (BFI) [38]. In their study, Bear and Fedio discovered that all 18 BFI traits were elevated when comparing TLE patients with healthy controls [38]. However, the lack of specificity and largely interrelated scales rendered the BFI to be an inaccurate measure of TLE personality [39]. Nevertheless, the BFI was successful in shifting the focus in epilepsy research from psychopathology to changes in behaviour [40]. Using modern personality models, we did not find any differences in personality between TLE and non-TLE patients. This is consistent with other studies using the five-factor model [41–43], but inconsistent with two studies using different models of personality. Specifically, investigations using a German personality questionnaire measuring neuroticism, extraversion, organic psychosyndrome and addiction, have generated different results [44,45]. While one study showed TLE patients to have greater neuroticism score, both studies showed TLE patients to have elevated introversion scores [44,45]. Although the reasons for the discrepancy is not entirely clear, cultural factors are one possibility. Despite these contradictory findings, the modern assessment of personality between TLE and non-TLE groups using the five-factor model has generally shown no major differences. Our findings are consistent with the emerging view that systematic personality differences between TLE and non-TLE patients do not exist.

Differences in personality profiles between patients with left and right TLE has been a less investigated area. Our study showed no differences in personality profiles and laterality which is consistent with a previous study in the literature using the same five-factor model [41]. In contrast, two other studies in the literature have shown to contradict these findings [38,46]. These studies found that patients with left TLE were more dependent and less composed, whereas patients with right TLE were found to have the tendency to underestimate their own problems. However, these studies were not performed using the five-factor model making them difficult to be used as studies for comparison. Further research using modern personality theory, such as the five-factor model, is warranted.

The strengths of our study include the use of a large sample size, gold standard VEM diagnosis, and the exclusion of non-diagnostic and mixed groups of patients. This method of patient sampling produces a high level of certainty regarding classification and allows for well-defined phenotypes, greatly increasing the accuracy of the data. Our study also has an advantage over most in the area of statistical analysis. The use of Bayesian statistics allows us to overcome the limitations of null hypothesis significance testing which has come under considerable criticism [47]. The built-in power analysis enables us to gather evidence for the null. For example, in our study, the domain agreeableness was revealed to support the null hypothesis, indicating that there are no differences in agreeableness between ES and PNES patients. To our knowledge, our study has also been the first to directly investigate the classification accuracy of the five-factor model in a clinical setting. While conventional statistical methods are used to identify group differences no matter how small, they do not address the clinically important question whether they assist in diagnosis. This is important to consider as often group differences and clinical classification are not aligned. Our study demonstrated considerable group differences present in the domain of openness and facet of aesthetic interests, however, they did not perform sufficiently well to facilitate clinical use of the NEO-FFI for diagnostic purposes.

The main limitation of our study is the absence of other clinical information (for example, the number of years since first seizure-like event, length and frequency of seizures, and anti-epileptic drugs (AEDs)) in the collected patient data. Some of these variables have been used in combination with other psychometric testing in the past to increase PNES classification accuracy [15,48,49]. However, this study was a clinical audit and it was necessary to limit the investigation to personality data and limited demographics. Further research should investigate the incremental validity of using other commonly collected variables in combination with the NEO-FFI. Our research study also has the same limitation as most other ES and PNES studies. Sampling from a tertiary referral population often results in a patient group with the most severe forms of the disease and may cause results to be heavily skewed. However, this sampling strategy is difficult to modify given the trade-off between a more representative population and a reduced accuracy of diagnosis.

The novel finding of elevated openness and aesthetic interests in PNES patients compared to ES patients may provide a new avenue for management strategies in PNES patients. Currently, management in PNES patients involves a mixture of antidepressants and psychotherapy [50,51]. Evidence for these strategies is limited and remains a large gap in the literature. Patients who tend to score higher on the domain of openness have been shown to be more receptive to unconventional forms of therapy such as imagery techniques [16]. Whilst no studies have investigated the relation between the facet of aesthetic interests and therapy, people who score higher on this facet have a greater appreciation for art and may be more suited to creative therapeutic modalities, such as art and music therapy. To our knowledge, there is no research studying the benefits of art and music therapy in PNES patients, however this presents a direction for future research. In addition, the lower extraversion in ES and PNES patients compared to the general population indicate that they may require more directed forms of therapy and antidepressant medication compared to patients who have high extraversion scores [16].

## 5. Conclusion

In conclusion, our study has been the first to investigate the use of modern models of personality to classify PNES and ES. While openness and aesthetic interests were demonstrated to be elevated in PNES patients, sensitivity and specificity were poor for diagnostic or screening purposes. Although the differences in response profiles were not sufficient for clinical classification, our study demonstrates that there is evidence for an association between personality profiles and these diagnostic groups. Repeating this study using more recent psychometric tests such as the NEO-Personality Inventory 3 or the Personality Inventory for DSM-5 is an avenue for new research.

## Data Availability

All data are available at http://doi.org/10.17605/OSF.IO/T3A9G.

## Abbreviations

AED: Anti-epileptic drugs
AUC: Area under the curve
BPPV: Benign paroxysmal positional vertigo
BFI: Bear-Fedio Inventory
ES: Epileptic seizures
Extra-TLE: Extra-Temporal lobe epilepsy
FFM: Five-factor model
ILAE: International League Against Epilepsy
MMPI: Minnesota Multiphasic Personality Inventory
NEO-FFI: Neo Five-Factor Inventory
PAI: Personality Assessment Inventory
PNES: Psychogenic non-epileptic seizures
PTSD: Post-traumatic stress disorder
TLE: Temporal Lobe epilepsy
RMH: Royal Melbourne Hospital
ROC: Receiver operating characteristic
VEM: Video-EEG monitoring

## Acknowledgements

We would like to thank all patients admitted to the Royal Melbourne Hospital’s VEM unit who have taken the time to participate in this study. We also would like to extend our appreciation to all staff who have assisted in the collection of data.

## Appendices

**Table A.1.** Personality domain descriptions

| *Domains* | *High* | *Low* |
| --- | --- | --- |
| Neuroticism | Emotional stability<br>Maladjustment<br>Tendency to experience negative affects<br>Impulsive<br>Poor coping mechanisms | Calm<br>Emotionally stable<br>Even-tempered |
| Extraversion | Sociable<br>Energetic<br>Assertive<br>Enjoys large gatherings | Reserved<br>Independent<br>Prefer solitude |
| Openness | Active imagination<br>Novel ideas<br>Unconventional values | Conservative<br>Conventional<br>Muted emotional responses |
| Agreeableness | Sympathetic<br>Eager to help others | Egocentric<br>Competitive<br>Distrustful |
| Conscientiousness | Strong-willed<br>Determined<br>Reliable<br>Punctual | Lackadaisical |

**Table A.2.** NEO-FFI domains and facets

| *Domains* | *Facets* |
| --- | --- |
| Neuroticism | Negative affect<br>Self-reproach |
| Extravertisism | Positive affect<br>Sociability<br>Activity |
| Openness | Aesthetic interests<br>Intellectual interests<br>Unconventionality |
| Agreeableness | Nonantagonistic orientation<br>Prosocial orientation |
| Conscientiousness | Orderliness<br>Goal striving<br>Dependability |

## References

[1] LaFrance Jr WC, Baker GA, Duncan R, Goldstein LH, Reuber M. Minimum requirements for the diagnosis of psychogenic nonepileptic seizures: a staged approach: a report from the International League Against Epilepsy Nonepileptic Seizures Task Force. Epilepsia 2013;54:2005–18.

[2] de Timary P, Fouchet P, Sylin M, Indriets JP, de Barsy T, Lefèbvre A, et al. Non-epileptic seizures: delayed diagnosis in patients presenting with electroencephalographic (EEG) or clinical signs of epileptic seizures. Seizure 2002;11:193–7.

[3] Magee JA, Burke T, Delanty N, Pender N, Fortune GM. The economic cost of nonepileptic attack disorder in Ireland. Epilepsy Behav 2014;33:45–8.

[4] Nowack WJ. Epilepsy: a costly misdiagnosis. Clin Electroencephalogr 1997;28:225–8.

[5] Whitaker JN. The confluence of quality of care, cost-effectiveness, pragmatism, and medical ethics in the diagnosis of nonepileptic seizures: a provocative situation for neurology. Arch Neurol 2001;58:2066–7.

[6] Karterud HN, Knizek BL, Nakken KO. Changing the diagnosis from epilepsy to PNES: Patients’ experiences and understanding of their new diagnosis. Seizure 2010;19:40–6.

[7] Benbadis SR. The EEG in nonepileptic seizures. J Clin Neurophysiol 2006:340–52.

[8] King DW, Gallagher BB, Murvin AJ, Smith DB, Marcus DJ, Hartlage LC, et al. Pseudoseizures: diagnostic evaluation. Neurology 1982;32:18–23.

[9] Brown RJ, Syed TU, Benbadis S, LaFrance Jr WC, Reuber M. Psychogenic nonepileptic seizures. Epilepsy Behav 2011;22:85–93.

[10] Iriarte J, Parra J, Urrestarazu E, Kuyk J. Controversies in the diagnosis and management of psychogenic pseudoseizures. Epilepsy Behav 2003;4:354–9.

[11] Ghougassian DF, d’Souza W, Cook MJ, O’Brien TJ. Evaluating the utility of inpatient video-EEG monitoring. Epilepsia 2004;45:928–32. doi:10.1111/j.0013-9580.2004.51003.x.

[12] Lagerlund TD, Cascino GD, Cicora KM, Sharbrough FW. Long-term electroencephalographic monitoring for diagnosis and management of seizures. Mayo Clin Proc 1996;71:1000–6. doi:10.1016/S0025-6196(11)63776-2.

[13] Watemberg N, Tziperman B, Dabby R, Hasan M, Zehavi L, Lerman-Sagie T. Adding video recording increases the diagnostic yield of routine electroencephalograms in children with frequent paroxysmal events. Epilepsia 2005;46:716–9. doi:10.1111/j.1528-1167.2005.50004.x.

[14] Cragar DE, Berry DT, Fakhoury TA, Cibula JE, Schmitt FA. A review of diagnostic techniques in the differential diagnosis of epileptic and nonepileptic seizures. Neuropsychol Rev 2002;12:31–64.

[15] Hill SW, Gale SD. Predicting psychogenic nonepileptic seizures with the Personality Assessment Inventory and seizure variables. Epilepsy Behav EB 2011;22:505–10. doi:10.1016/j.yebeh.2011.08.001.

[16] Costa PT, MacCrae RR. Revised NEO personality inventory (NEO PI-R) and NEO five-factor inventory (NEO-FFI): Professional manual. Psychological Assessment Resources, Incorporated; 1992.

[17] Cragar D.E., Berry D.T.R., Schmitt F.A., Fakhoury T.A. Cluster analysis of normal personality traits in patients with psychogenic nonepileptic seizures. Epilepsy Behav 2005;6:593–600. doi:10.1016/j.yebeh.2005.03.007.

[18] Vilyte G, Pretorius C. Personality traits, illness behaviors, and psychiatric comorbidity in individuals with psychogenic nonepileptic seizures (PNES), epilepsy, and other nonepileptic seizures (oNES): Differentiating between the conditions. Epilepsy Behav 2019;98:210–9.

[19] Fisher RS, Boas WVE, Blume W, Elger C, Genton P, Lee P, et al. Epileptic seizures and epilepsy: definitions proposed by the International League Against Epilepsy (ILAE) and the International Bureau for Epilepsy (IBE). Epilepsia 2005;46:470–2.

[20] Fisher RS, Acevedo C, Arzimanoglou A, Bogacz A, Cross JH, Elger CE, et al. ILAE official report: a practical clinical definition of epilepsy. Epilepsia 2014;55:475–82.

[21] Association AP. Diagnostic and statistical manual of mental disorders (DSM-5®). American Psychiatric Pub; 2013.

[22] Berg AT, Berkovic SF, Brodie MJ, Buchhalter J, Cross JH, van Emde Boas W, et al. Revised terminology and concepts for organization of seizures and epilepsies: report of the ILAE Commission on Classification and Terminology, 2005–2009. Epilepsia 2010;51:676–85.

[23] John O. The “Big Five” factor taxonomy: Dimensions of personality in the natural language and in questionnaires. Handb. Personal. Res., 1990, p. 66–100.

[24] McCrae RR. The five-factor model and its assessment in clinical settings. J Pers Assess 1991;57:399–314. doi:10.1207/s15327752jpa5703_2.

[25] Digman J. Personality structure:Emergence of the five-factor model. Annu Rev Clin Psychol 1990;41:417–40.

[26] McCrae RR. NEO-PIR data from 36 cultures. Five-Factor Model Personal. Cult., Springer; 2002, p. 105–25.

[27] Saucier G. Replicable item-cluster subcomponents in the NEO Five-Factor Inventory. J Pers Assess 1998;70:263–76.

[28] JASP Team. JASP(Version 0.9.2)[Computer software]. 2019.

[29] R Core Team. R: A Language and Environment for Statistical Computing. 2019.

[30] Cohen J. Statistical power analysis for the behavioral sciences. Routledge; 2013.

[31] DeYoung CG. Openness/Intellect: A dimension of personality reflecting cognitive exploration. APA Handb Personal Soc Psychol Personal Process Individ Differ 2014;4:369–99.

[32] Zillig LMP, Hemenover SH, Dienstbier RA. What do we assess when we assess a Big 5 trait? A content analysis of the affective, behavioral, and cognitive processes represented in Big 5 personality inventories. Pers Soc Psychol Bull 2002;28:847–58.

[33] Elger CE, Helmstaedter C, Kurthen M. Chronic epilepsy and cognition. Lancet Neurol 2004;3:663–72.

[34] Ficker DM, O’Brien TJ. Epilepsy and the interictal state: co-morbidities and quality of life. John Wiley & Sons; 2015.

[35] Blumer D. Evidence supporting the temporal lobe epilepsy personality syndrome. Neurology 1999;53:S9–12.

[36] Waxman SG, Geschwind N. The interictal behavior syndrome of temporal lobe epilepsy. Arch Gen Psychiatry 1975;32:1580–6.

[37] Geschwind N. Behavioural changes in temporal lobe epilepsy. Psychol Med 1979;9:217–9.

[38] Bear DM, Fedio P. Quantitative analysis of interictal behavior in temporal lobe epilepsy. Arch Neurol 1977;34:454–67.

[39] Sorensen AS, Bolwig TG. Personality and epilepsy: new evidence for a relationship? A review. Compr Psychiatry 1987;28:369–83.

[40] Devinsky O, Najjar S. Evidence against the existence of a temporal lobe epilepsy personality syndrome. Neurology 1999;53:S13–25.

[41] Swinkels WAM, Van Emde Boas W, Kuyk J, Van Dyck R, Spinhoven P. Interictal Depression, Anxiety, Personality Traits, and Psychological Dissociation in Patients with Temporal Lobe Epilepsy (TLE) and Extra-TLE: INTERICTAL PSYCHOPATHOLOGY IN TLE AND EXTRA-TLE. Epilepsia 2006;47:2092–103. doi:10.1111/j.1528-1167.2006.00808.x.

[42] Locke DEC, Fakhoury TA, Berry DTR, Locke TR, Schmitt FA. Objective evaluation of personality and psychopathology in temporal lobe versus extratemporal lobe epilepsy. Epilepsy Behav 2010;17:172–7. doi:10.1016/j.yebeh.2009.11.005.

[43] Pung T, Schmitz B. Circadian rhythm and personality profile in juvenile myoclonic epilepsy. Epilepsia 2006;47:111–4.

[44] Witt J-A, Hollmann K, Helmstaedter C. The impact of lesions and epilepsy on personality and mood in patients with symptomatic epilepsy: a pre-to postoperative follow-up study. Epilepsy Res 2008;82:139–46. doi:10.1016/j.eplepsyres.2008.07.011.

[45] Helmstaedter C, Witt J-A. Multifactorial etiology of interictal behavior in frontal and temporal lobe epilepsy. Epilepsia 2012;53:1765–73. doi:10.1111/j.1528-1167.2012.03602.x.

[46] Feddersen B, Herzer R, Hartmann U, Gaab MR, Runge U. On the psychopathology of unilateral temporal lobe epilepsy. Epilepsy Behav EB 2005;6:43–9.

[47] Wasserstein RL, Lazar NA. The ASA’s statement on p-values: context, process, and purpose. Am Stat 2016;70:129–33.

[48] Storzbach D., Binder L.M., Salinsky M.C., Campbell B.R., Mueller R.M. Improved prediction of nonepileptic seizures with combined MMPI and EEG measures. Epilepsia 2000;41:332–7. doi:10.1111/j.1528-1157.2000.tb00164.x.

[49] Schramke C.J., Valeri A., Valeriano J.P., Kelly K.M. Using the Minnesota Multiphasic Inventory 2, EEGs, and clinical data to predict nonepileptic events. Epilepsy Behav 2007;11:343–6. doi:10.1016/j.yebeh.2007.06.011.

[50] LaFrance WC, Keitner GI, Papandonatos GD, Blum AS, Machan JT, Ryan CE, et al. Pilot pharmacologic randomized controlled trial for psychogenic nonepileptic seizures. Neurology 2010;75:1166–73. doi:10.1212/WNL.0b013e3181f4d5a9.

[51] Goldstein LH, Chalder T, Chigwedere C, Khondoker MR, Moriarty J, Toone BK, et al. Cognitive-behavioral therapy for psychogenic nonepileptic seizures: a pilot RCT. Neurology 2010;74:1986–94. doi:10.1212/WNL.0b013e3181e39658.

